# Comprehensive aortic stenosis characterization using multi-view deep learning

**DOI:** 10.1101/2025.09.26.25336778

**Authors:** Hirotaka Ieki, Yuki Sahashi, Miloš Vukadinovic, Meenal Rawlani, Christina Binder, Neal Yuan, Andrew P. Ambrosy, Alan S. Go, Wansu Chen, Ming-Sum Lee, Bryan He, Paul Cheng, David Ouyang

## Abstract

**Background and Aims:** Accurate assessment of aortic stenosis (AS) requires integration of both structural and functional information characterized by visual traits as well as quantitation of gradients. Existing artificial intelligence (AI) models utilize solely either structural or functional information.

**Methods:** We developed EchoNet-AS, an open-source end-to-end integrated approach combining video based convolutional neural networks to assess valve motion as well as segmentation models to automate the measurement of aortic valve peak velocity to classify AS severity.

**Results:** EchoNet-AS was trained on 210,193 images from 16,076 studies from Kaiser Permanente Northern California (KPNC) and validated on 1,589 held-out test studies and a temporally distinct cohort of 19,206 studies. The final model was also externally validated on 2,415 studies from Stanford Healthcare (SHC) and 9,038 studies from Cedars-Sinai Medical Center (CSMC). Combining assessments from multiple echocardiographic videos and Doppler measurements, EchoNet-AS achieved excellent discrimination of severe AS with AUC 0.964 [95% CI: 0.952 – 0.973] in the KPNC held-out cohort and 0.985 [0.981 – 0.988] in the temporally distinct cohort, which was superior to models using single views or only Doppler measurements. The performance was consistently robust in distinct external cohorts with an AUC 0.985 [0.975 – 0.992] at SHC and 0.989 [0.986 – 0.992] at CSMC.

**Conclusions:** EchoNet-AS synthesizes information from both B-mode videos and Doppler images to accurately assess AS severity. Its strong performance generalizes robustly to external validation cohorts and shows potential as an automated clinical decision support tool.

## Introduction

Aortic stenosis (AS) is the most common heart valve disease, especially among older adults, and is associated with high rate of mortality and morbidity^1–3^. Echocardiography is the primary modality for assessing AS severity, and with the advent of transcatheter aortic valve replacement (TAVR), accurate evaluation of AS has become increasingly critical for identifying patients who require intervention, tailoring monitoring frequency for disease progression, and guiding treatment decisions. However, the current diagnostic standard of using human-interpreted transthoracic echocardiograms is limited by interobserver variability and the subjective nature of video interpretation. This is particularly problematic at the clinically important moderate–severe boundary, where misclassification can lead to delayed TAVR referral, unnecessary repeat echocardiograms, or inconsistent treatment recommendations across sites.

Recent advances in deep learning methods have enabled severity prediction of valvular heart disease from echocardiography images or videos^4–9^. For AS, prior studies have demonstrated that artificial intelligence (AI) models can accurately detect severe AS using single-view videos^10,11^, and others have proposed a commercialized software to automate Doppler-derived measurements to assess AS severity^12,13^. However, in clinical practice, it is important to review multiple videos and account for physiologic factors (especially in more challenging diagnostic scenarios such as with low-flow low-gradient AS) to holistically understand the severity of AS^14,15^.

Given that existing approaches do not leverage the complementary information available from multiple videos and Doppler measurements in a comprehensive echocardiographic assessment, we hypothesized that an integrated framework combining all relevant echocardiographic information could improve the accuracy and clinical reliability of AS assessment. We developed an open-source deep learning framework, EchoNet-AS, which integrates information from multiple echocardiographic views and Doppler measurements to predict the severity of AS (**Figure. 1**) and validated the framework across multiple institutions to demonstrate its generalizability.

**Figure 1.**
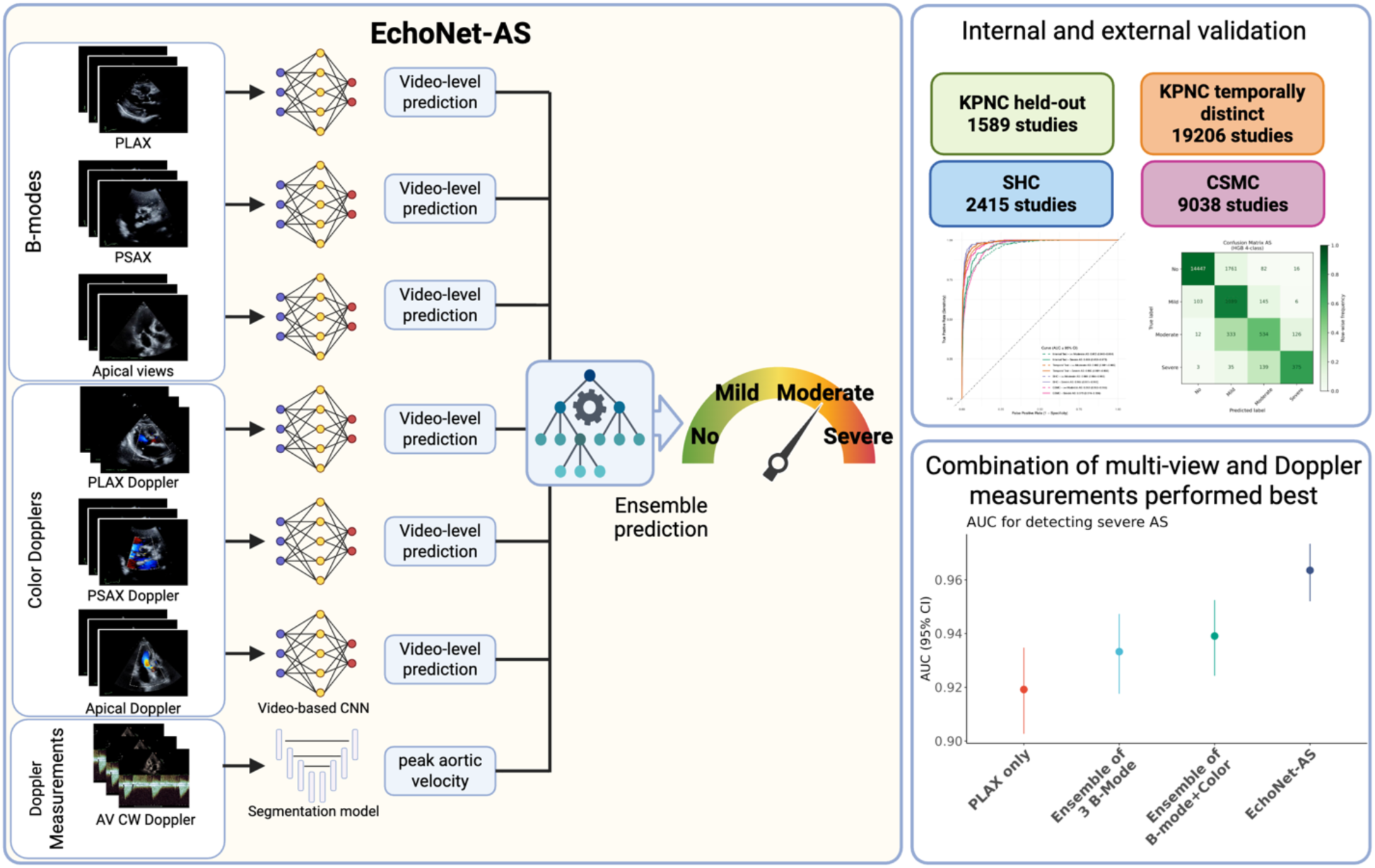
Overview of the study. In the EchoNet-AS framework, we trained video-based convolutional neural network (CNN) models to predict the severity of aortic stenosis (AS) from six distinct transthoracic echocardiographic views including both B-modes [parasternal long axis (PLAX), parasternal short axis (PSAX) and apical five/three chamber views] and Color dopplers [PLAX Doppler, PSAX Doppler and Apical Doppler]. In parallel, a segmentation model automatically measures peak aortic jet velocity. These outputs were integrated through an ensemble approach to generate a final prediction of AS severity. We evaluated the performance of this framework across four independent test datasets including held-out cohort and temporally distinct cohort from Kaiser Permanente Northern California (KPNC) and external validation cohort from Stanford Healthcare (SHC) and Cedars-Sinai Medical Center (CSMC). EchoNet-AS, which integrates B-modes, color Dopplers and Doppler measurements, showed superior performance compared to the single-view model or video only models. Created in https://BioRender.com

## Methods

### Study population, imaging and data source

In this multicenter retrospective study, we used transthoracic echocardiography (TTE) data from three different medical centers: Kaiser Permanente Northern California (KPNC), Stanford Healthcare (SHC), and Cedars-Sinai Medical Center (CSMC) (**Figure 2**). A total of 608,086 TTE videos from 48,324 studies, obtained from 45,852 unique patients, were used for model development and testing. In KPNC, 17,665 studies conducted between January 1, 2022 and December 31, 2023 were sampled, with cases without AS downsampled to mitigate class imbalance during model training. This cohort was split into a derivation dataset (= training + validation; 16,076 TTE studies from 15,213 patients) and a held-out test cohort (1,589 TTE studies from 1,583 patients), ensuring no patient overlap between the subsets. The final derivation dataset comprised of 16,076 studies with 210,193 echocardiographic videos.

**Figure 2.**
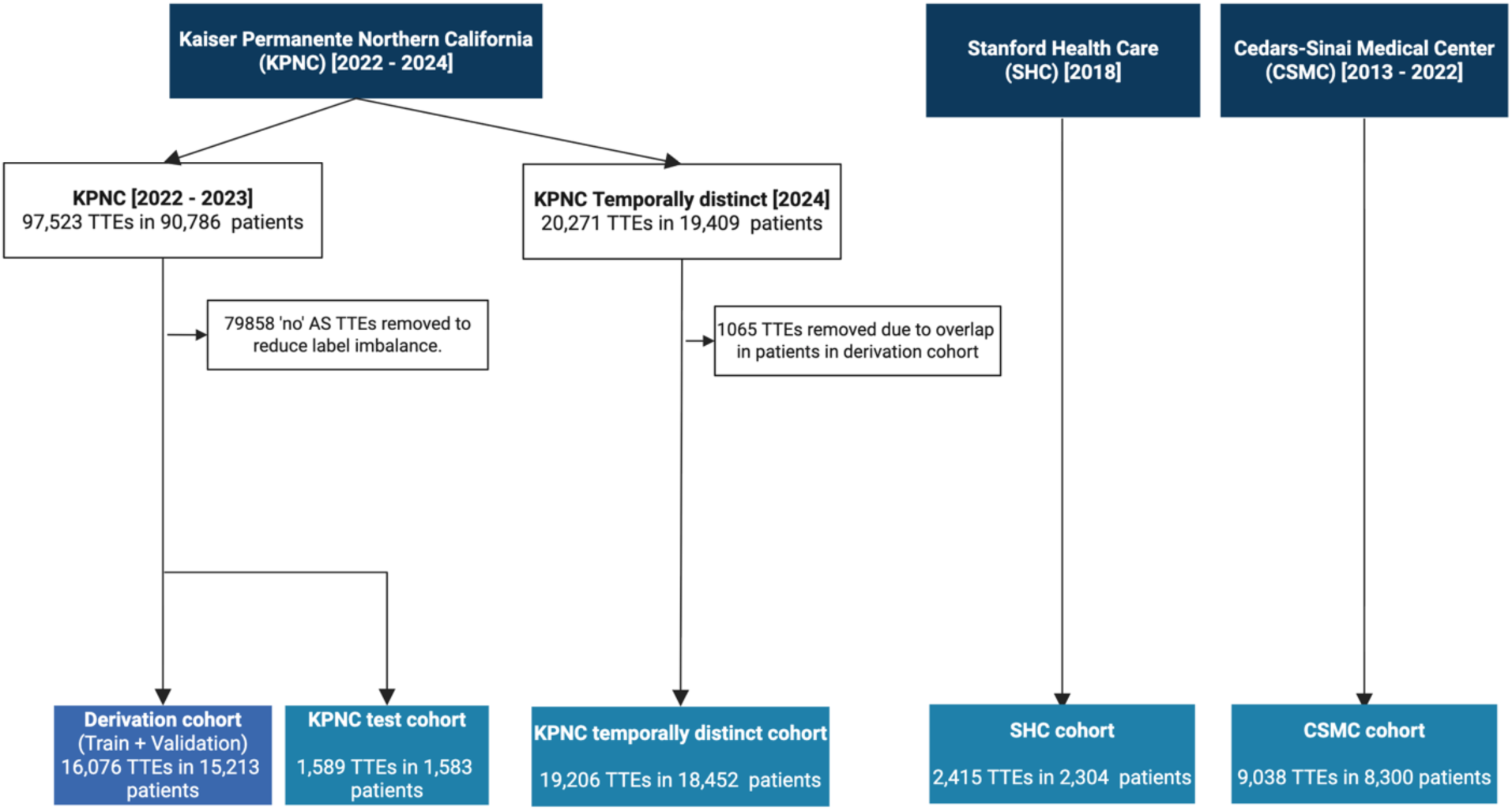
Data flowchart The construction of the study cohorts from echocardiographic databases from three different medical centers: Kaiser Permanente Northern California (KPNC), Stanford Health Care (SHC), and Cedars-Sinai Medical Center (CSMC). From the KPNC database, 97,523 TTEs from 90,786 patients (2022–2023) were filtered to form a balanced cohort of 17,665 TTEs in 16,796 patients by removing 79,858 “none or trace“ AS severity studies. This cohort was split into a derivation dataset (16,076 TTEs for traing and validation data) and an internal test cohort (1,589 TTEs), ensuring no patient overlap between the subsets. Separately, 20,271 TTEs from 2024 were obtained, and after excluding 1,065 overlapping patients’ TTEs with the derivation cohort, 19,206 TTEs from 18,452 patients were used as the temporally distinct test cohort. From the Stanford and Cedars-Sinai databases, external validation cohorts were formed with 2,415 TTEs from 2,304 patients and 9,038 TTEs from 8,300 patients, respectively. Created in https://BioRender.com

To assess temporal generalizability, we additionally collected 20,271 TTE studies conducted in 2024, of which 19,206 studies from 18,452 unique patients remained after excluding overlapping individuals from the derivation dataset. All the videos were recorded from adult participants on Philips (Amsterdam, The Netherlands) EPIQ 7C, EPIQ CVx, Affiniti 70C or Affiniti CVx machines. The acquisition of images followed a comprehensive and standardized protocol in-line with current recommendations^14,16,17^. The images were stored as Digital Imaging and Communications in Medicine (DICOM) files, and these files underwent pre-processing to ensure consistency and extraneous details beyond the ultrasound section were removed, and metadata was de-identified.

Additionally, our model was evaluated on two distinct external cohorts from separate healthcare systems. A total of 2,415 studies from 2,304 patients conducted in 2018 were extracted from Stanford Healthcare (SHC; Stanford California, USA) database. In addition, 9,038 TTEs from 8,300 patients conducted between 2013 and 2022 at the Cedars-Sinai Medical Center (CSMC; Los Angeles, California, USA) were extracted. The SHC and CSMC echocardiography lab uses Philips EPIQ 7 or IE33 machines and follows a similar standardized comprehensive protocol. AS severity was extracted from finalized echo report. Finally, to compare our model with the previously published model to detect severe AS, we extracted the same 5,572 TTE studies from CSMC, and then followed the approach of a prior study with a cohort of 4,226 studies for head-to-head comparison (CSMC replication cohort)^11^. For all cohorts, AS severity was categorized as ‘no, ‘mild’, mild to moderate’, ‘moderate’, ‘moderate to severe’ or ‘severe’. The reference labels used for training and evaluation were derived directly from the finalized physician echocardiography reports. No additional adjudication or consensus review was performed as part of this study, reflecting real-world reporting practice.

### AI model development

We developed a two-stage framework for AS assessment, termed EchoNet-AS, consisting of video-based deep learning predictions from both B-mode/Doppler videos and ensemble integration with the model outputs and Doppler measurements (**Figure 1**). In the first stage, six separate video-based convolutional neural networks (CNNs) based on the ResNet R(2+1)D architecture^18^, which has demonstrated strong performance in echocardiography-related video tasks^4^. These models were trained to classify AS severity from six standard echocardiographic views: parasternal long axis (PLAX), parasternal short axis (PSAX), apical five/three chamber views (AP), PLAX color Doppler, PSAX color Doppler, and AP color Doppler. The input videos were processed to mask the embedded texts. The resulting videos were subsampled by one in every two frames to yield 16 frames, and resized to 112 × 112 pixel. Cross-entropy loss was employed as the loss function, and early stopping after 10 epoch based on validation area under the receiver operating characteristic curve (AUC) were employed to prevent overfitting. All the models were trained using the Pytorch 2.5.0 and PyTorch Lightning 2.5.1 ^19^ deep learning framework with the Rectified Adam Schedule Free optimizer ^20^ with a batch size of 64 and mixed-precision training (bfloat16) ^21^. Training was performed on NVIDIA L40S GPUs, and the model weights from the epoch achieving the highest validation AUC were selected as the final models. In parallel, we utilized a previously described segmentation model based on DeepLabv3 (EchoNet-Measurement) that enables automated estimation of aortic peak velocity, achieving high accuracy with a mean absolute error of 10.4 cm/s in external validation^22^.

The second stage of the framework consisted of an ensemble model. The outputs from six view-specific networks—each generating a probability vector across the six AS severity classes— together with the peak aortic velocity measurement were used as input features for a machine learning ensemble implemented with the HistGradientBoostingClassifier in scikit-learn^23^. The final output is the probabilities of four AS severity classes (‘no’, ‘mild’, ‘moderate’ or ‘severe’). When multiple videos were available for the same view, the model outputs were averaged prior to integration. The ensemble classifier is robust to missing values, thereby allowing inclusion of studies that did not contain all six views. In parallel, we also constructed ensembles using only the three B-mode models as well as ensembles incorporating all six models (the three B-mode models and the three color-Doppler models). Hyperparameter tuning was performed using the validation dataset.

### Statistical analysis

The EchoNet-AS framework was evaluated across four independent test cohorts. For the description of the baseline cohort characteristics, categorical variables are presented as counts and percentages, while continuous variables are expressed as mean and standard deviation. We calculated the area under the receiver operating characteristic curve (AUC) as the main outcome. Positive predictive value (PPV), negative predictive value (NPV), recall (sensitivity), specificity and F1 score were calculated for clinically significant AS, which was defined as greater than moderate AS and severe AS. Intermediate AS categories (e.g. “mild to moderate”) were upgraded to the upper categories (e.g. “moderate”). All 95% confidence intervals were calculated from 10,000 bootstrapping replicates. Subgroup analysis was conducted in KPNC temporally distinct cohort and CSMC cohort to assess model performance in patients with different ranges of age, sex, body mass index (BMI), left ventricular ejection fraction (LVEF), associated comorbidities (at least moderate aortic regurgitation, mitral regurgitation, mitral stenosis, and tricuspid regurgitation), aortic valve morphology, and other clinical characteristics. To understand which regions of the echocardiographic videos contributed most for AI model assessment, we employed smooth grad saliency mapping^24^. For ensemble model, we calculated the feature importance of input features by permutation importance function in scikit-learn and summarized them by echocardiographic view level. Analyses were performed using Python (version 3.12.3) and R (version 4.4.3)

## Results

### Study population

The KPNC cohort patients (mean age: 73.1 ± 14.2 years in derivation cohort, 72.3 ± 14.8 years in held-out test cohort, 74.0 ± 14.2 years in temporally distinct test cohort) were older than external validation cohorts (65.7 ± 16.2 years in the SHC cohort and 65,7 ± 16.8 years in the CSMC cohort). The KPNC derivation and held-out test cohort were enriched for mild/moderate/severe AS patients to mitigate class imbalance during model training (see Method). The proportion of male participants, racial/ethnic groups, the mean left ventricular ejection fraction, and major comorbidities were similar across datasets. A detailed description of the characteristics of the cohort is shown in **Table 1**.

**Table 1.** Table of baseline demographic and echocardiographic characteristics in each cohort.

|  | Training<br>+Validation | Test data |  |  |  |
| --- | --- | --- | --- | --- | --- |
|  | KPNC Derivation<br>Dataset | KPNC held-out<br>test Cohort | KPNC temporally<br>distinct Cohort | SHC cohort | CSMC cohort |
| Number of studies | 16076 | 1589 | 19206 | 2415 | 9038 |
| Number of unique patients | 15213 | 1583 | 18452 | 2304 | 8300 |
| Number of videos | 210193 | 9534 | 248738 | 26757 | 112864 |
| Age, mean (SD), years | 73.1 (14.2) | 72.3 (14.8) | 74.0 (11.5) | 65.7 (16.2) | 65.7 (16.8) |
| Male sex, n (%) | 8326 (51.8) | 837 (52.7) | 9817 (51.1) | 1272 (52.7) | 4966 (54.9) |
| Race, n (%) |  |  |  |  |  |
| Asian | 2069 (12.9) | 212 (13.3) | 2088 (10.9) | 412 (17.1) | 652 (7.2) |
| Black | 997 (6.2) | 94 (5.9) | 2142 (11.2) | 141 (5.8) | 1272 (14.1) |
| Native American | 62 (0.4) | 7 (0.4) | 69 (0.4) | 8 (0.3) | 27 (0.3) |
| Pacific Islander | 113 (0.7) | 12 (0.8) | 82 (0.4) | 35 (1.4) | 31 (0.3) |
| White | 10280 (63.9) | 1015 (63.9) | 12345 (64.3) | 1361 (56.4) | 6204 (68.6) |
| Other/Unknown | 2555 (15.9) | 249 (15.7) | 2480 (12.9) | 458 (19.0) | 852 (9.4) |
| Hispanic ethnicity, n (%) | 2427 (15.1) | 238 (15.0) | 2335 (12.2) | 234 (9.7) | 1184 (13.1) |
| Height, mean (SD), cm | 168.2 (10.9) | 168.2 (10.9) | 169.3 (10.8) | 167.8 (11.4) | 168.9 (11.0) |
| Weight, mean (SD), kg | 82.7 (22.9) | 82.7 (23.0) | 83.2 (23.5) | 77.8 (21.5) | 77.0 (20.8) |
| BMI, mean (SD), kg/m^2 | 29.1 (7.1) | 29.1 (7.1) | 28.9 (7.1) | 27.0 (6.5) | 26.9 (6.4) |
| BSA, mean (SD), m^2 | 2.0 (0.3) | 2.0 (0.3) | 2.0 (0.3) | 1.9 (0.3) | 1.9 (0.3) |
| Hypertension, n (%) | 11177 (69.5) | 1077 (67.8) | 13385 (69.7) | 1328 (55.0) | 1673 (18.5) |
| Atrial fibrillation, n (%) | 4720 (29.4) | 451 (28.4) | 6446 (33.6) | 845 (35.0) | 747 (8.3) |
| History of aortic valve surgery, n (%) | 280 (1.7) | 22 (1.4) | 312 (1.6) | 2 (0.1) | 11 (0.1) |
| Bicuspid AV, n (%) | 456 (2.8) | 57 (3.6) | 80 (0.4) | 46 (1.9) | 112 (1.2) |
| AS severity, n (%) |  |  |  |  |  |
| no/trace | 4874 (30.3) | 547 (34.4) | 16306 (84.9) | 1982 (82.1) | 8283 (91.6) |
| mild | 4925 (30.6) | 489 (30.8) | 1343 (7.0) | 219 (9.1) | 239 (2.6) |
| moderate | 3718 (23.1) | 346 (21.8) | 1005 (5.2) | 104 (4.3) | 238 (2.6) |
| severe | 2559 (15.9) | 207 (13.0) | 552 (2.9) | 110 (4.6) | 278 (3.1) |
| AV peak velocity, mean (SD), m/s | 2.5 (1.0) | 2.5 (1.0) | 1.7 (0.7) | 2.1 (1.0) | 1.6 (0.9) |
| AV mean gradient, mean (SD), mmHg | 17.1 (13.9) | 16.4 (13.8) | 7.2 (7.8) | 12.5 (14.2) | 9.2 (10.9) |
| AV area, mean (SD), cm^2 | 1.6 (0.7) | 1.6 (0.7) | 2.2 (0.7) | 2.0 (0.9) | 2.1 (0.8) |
| At least moderate AR, n (%) | 1285 (8.0) | 122 (7.7) | 917 (4.8) | 173 (7.2) | 562 (6.2) |
| At least moderate MS, n (%) | 378 (2.4) | 38 (2.4) | 157 (0.8) | 47 (1.9) | 55 (0.6) |
| At least moderate MR, n (%) | 1367 (8.5) | 123 (7.7) | 1567 (8.2) | 365 (15.1) | 1326 (14.7) |
| At least moderate TR, n (%) | 1485 (9.2) | 122 (7.7) | 1915 (10.0) | 418 (17.3) | 1540 (17.0) |
| IVSd, mean (SD), cm | 1.1 (0.3) | 1.1 (0.3) | 1.1 (0.2) | 1.1 (0.2) | 1.1 (0.3) |
| LVPWd, mean (SD), cm | 1.1 (0.2) | 1.1 (0.2) | 1.1 (0.2) | 1.0 (0.2) | 1.1 (0.2) |
| LVEF, mean (SD), % | 59.0 (9.6) | 59.2 (9.8) | 57.9 (10.3) | 57.7 (12.1) | 57.3 (14.5) |
| RVSP, mean (SD), mmHg | 28.1 (12.0) | 28.2 (11.8) | 27.7 (11.7) | 36.0 (14.8) | 31.6 (13.8) |

BMI, body mass index; BSA, body surface area; AV, aortic valve; AS, aortic stenosis; AR, aortic
regurgitation; MS, mitral stenosis; MR, mitral regurgitation; TR, tricuspid regurgitation; IVSd inter-
ventricular septum diastolic thickness; LVPWd, left ventricular posterior wall diastolic thickness;
LVEF, left ventricular ejection fraction; RVSP, right ventricular systolic pressure; KPNC, Kaiser
Permanente Northern California; SHC, Stanford Healthcare; CSMC, Cedars-Sinai Medical Center.

### Model performance

In the initial analysis of the performance of individual models for each standard echocardiographic view, the PLAX view model demonstrated the best performance in detecting severe AS (**sTable 1**). While performance varied across individual views, with the PLAX and apical views generally achieving the highest AUCs, the ensemble models that integrated multiple views consistently outperformed any single-view model across all cohorts (**Table 2**). In the KPNC temporally distinct test data, the ensemble of 3 B-mode views (PLAX, PSAX and AP) achieved an AUC of 0.976 [0.973 – 0.980] in the temporally distinct test data and the ensemble of all 6 video models achieved an AUC of 0.979 [0.975 – 0.983]. By incorporating automated Doppler measurements, the full EchoNet-AS framework demonstrated the best performance, with an AUC of 0.955 [0.946 - 0.964] for moderate or severe AS and 0.964 [0.952-0.973] for severe AS in the KPNC held-out test cohort and 0.983 [0.980 - 0.985] for moderate or severe AS and 0.985 [0.981-0.988] for severe AS in the KPNC temporally distinct cohort. The Echonet-AS generalized well in the external SHC (AUC, moderate or severe AS 0.989 [0.984 – 0.993], severe AS 0.985 [0.975 – 0.992]) and CSMC cohort (AUC; moderate or severe AS 0.978 [0.974 – 0.982]; severe AS 0.989 [0.986 – 0.992]) (**Table 2**, **Figure 3**).

**Figure 3.**
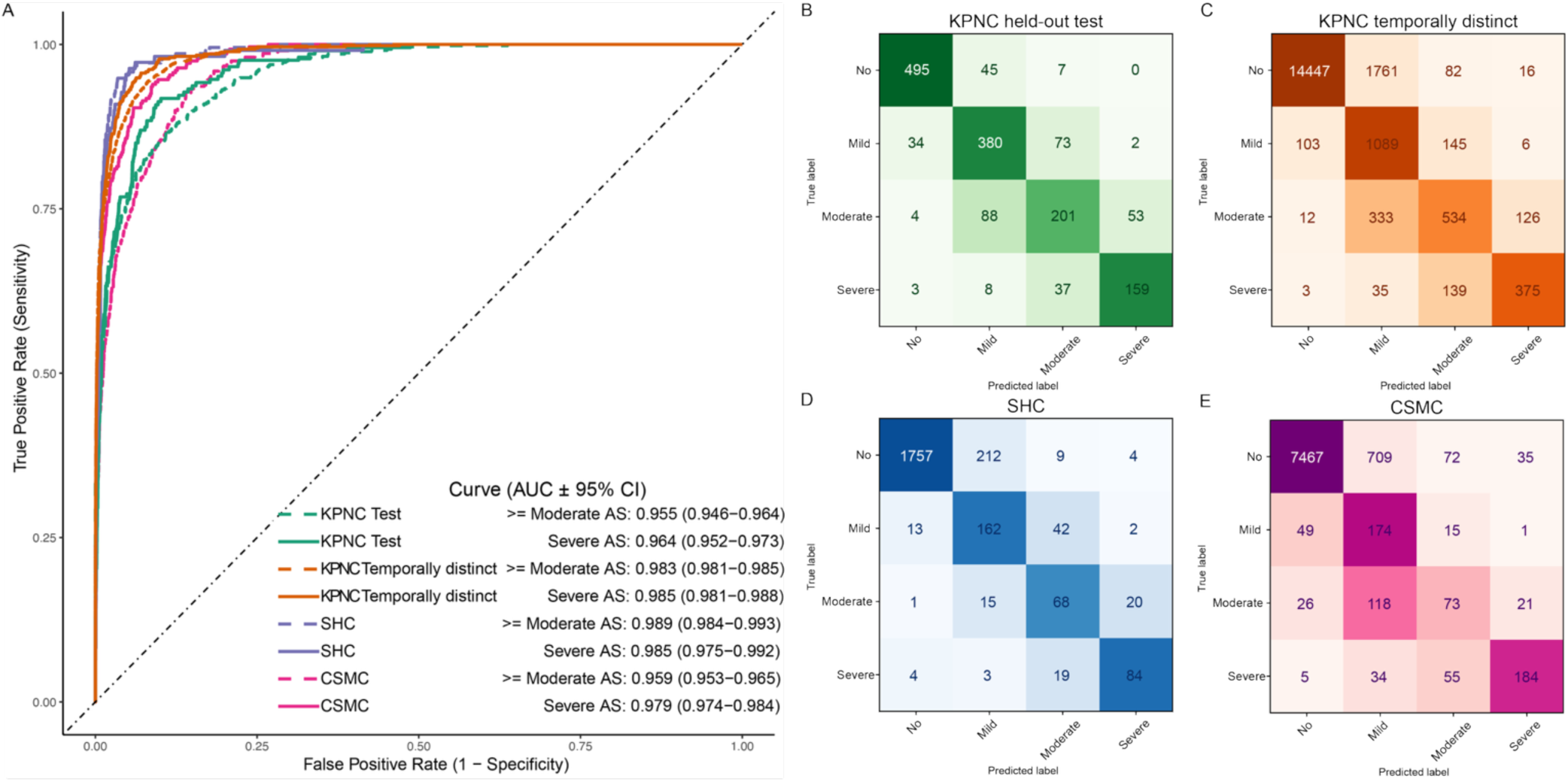
(A) Receiver operating characteristic (ROC) curves for detection of severe or at least moderate aortic stenosis (AS) in Kaiser Permanente Northern California (KPNC) internal test cohort (green), KPNC temporal cohort (orange), Stanford Healthcare (SHC) cohort (purple), and Cedars-Sinai Medical Center (CSMC) cohort (pink). Predictions for severe and at least moderate are shown in solid and dash lines, respectively. (B-E) Confusion matrix of AS severity classification on KPNC test cohort (B), KPNC temporally distinct cohort (C), SHC cohort (D), and CSMC cohort (E) are shown. Confusion matrix colormap values were scaled based on the proportion of actual disease cases in each class that were predicted in each possible disease category. AUC area under the receiver operating characteristic curve; CI, confidence interval.

**Table 2.** AUC of discriminating severe or moderate+ aortic stenosis in the test cohorts by different models.

| Models | Severe AS |  |  |  | ≥ Moderate AS |  |  |  |
| --- | --- | --- | --- | --- | --- | --- | --- | --- |
|  | KPNC Test | KPNC Temporally Distinct | SHC | CSMC | KPNC Test | KPNC Temporally Distinct | SHC | CSMC |
| EchoNet-AS | 0.964<br>(0.952 – 0.973) | 0.985<br>(0.981 – 0.988) | 0.985<br>(0.975 – 0.992) | 0.989<br>(0.986 – 0.992) | 0.955<br>(0.946 – 0.964) | 0.983<br>(0.980 – 0.985) | 0.989<br>(0.984 – 0.993) | 0.978<br>(0.974 – 0.982) |
| Ensemble of B Modes + Color Dopplers | 0.939<br>(0.924 – 0.952) | 0.979<br>(0.975 – 0.983) | 0.980<br>(0.971 – 0.988) | 0.985<br>(0.978 – 0.990) | 0.926<br>(0.914 – 0.938) | 0.976<br>(0.973 – 0.978) | 0.983<br>(0.977 – 0.988) | 0.977<br>(0.972 – 0.981) |
| Ensemble of B Modes | 0.933<br>(0.918 – 0.947) | 0.976<br>(0.973 – 0.980) | 0.973<br>(0.961 – 0.983) | 0.981<br>(0.974 – 0.986) | 0.915<br>(0.902 – 0.928) | 0.969<br>(0.966 – 0.972) | 0.979<br>(0.973 – 0.985) | 0.971<br>(0.965 – 0.976) |
| PLAX | 0.919<br>(0.903 – 0.935) | 0.970<br>(0.964 – 0.974) | 0.973<br>(0.964 – 0.982) | 0.972<br>(0.964 – 0.979) | 0.910<br>(0.895 – 0.923) | 0.967<br>(0.963 – 0.970) | 0.975<br>(0.968 – 0.982) | 0.968<br>(0.962 – 0.973) |

KPNC, Kaiser Permanente Northern California; SHC, Stanford Healthcare; CSMC, Cedars-Sinai
Medical Center; PLAX, parasternal long axis view; ≥ Moderate AS, moderate or severe AS

Across all cohorts, positive predictive value (PPV) ranged from 0.731 to 0.848, negative predictive value (NPV) from 0.909 to 0.991, recall from 0.622 to 0.911, specificity from 0.917 to 0.996, and F1-score from 0.701 to 0.848, indicating consistent performance across diverse evaluation metrics (**Table 3**). Finally, we compared EchoNet-AS with the published severe AS detection model. Since the model weights were not publicly available, we extracted the same 5,572 TTE studies from the CSMC database and downsampled severe-AS cases as described in the published paper^11^, yielding 4,226 studies including 65 severe AS cases. In this CSMC replicated cohort, our ensemble of multi-view and doppler measurement approach demonstrated better severe AS discrimination performance with AUC of 0.976 [0.970 – 0.979] (**sTable 2**). In the CSMC replication cohort (N=4,226), our single view model showed comparable performance compared to prior approaches (AUC 0.956 [0.940 – 0.969]) to detect severe AS, while the ensemble approach with EchoNet-AS yielded superior overall performance with AUC of 0.972 [0.963 – 0.979].

**Table 3.**
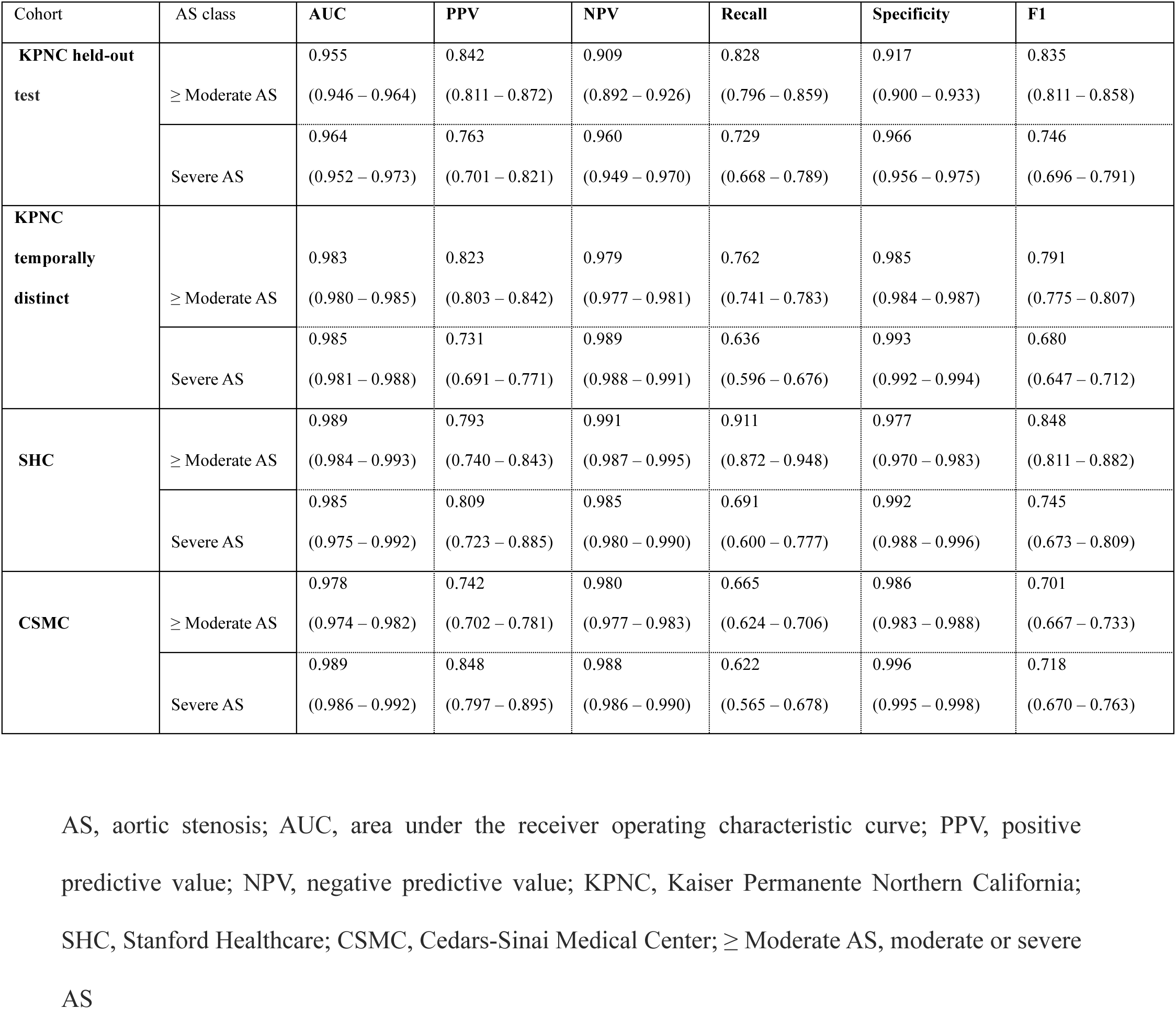
Different performance metrics of EchoNet-AS to detect severe or at least moderate aortic stenosis in the test cohorts.

| Cohort | AS class | AUC | PPV | NPV | Recall | Specificity | F1 |
| --- | --- | --- | --- | --- | --- | --- | --- |
| **KPNC held-out test** | ≥ Moderate AS | 0.955<br>(0.946 – 0.964) | 0.842<br>(0.811 – 0.872) | 0.909<br>(0.892 – 0.926) | 0.828<br>(0.796 – 0.859) | 0.917<br>(0.900 – 0.933) | 0.835<br>(0.811 – 0.858) |
|  | Severe AS | 0.964<br>(0.952 – 0.973) | 0.763<br>(0.701 – 0.821) | 0.960<br>(0.949 – 0.970) | 0.729<br>(0.668 – 0.789) | 0.966<br>(0.956 – 0.975) | 0.746<br>(0.696 – 0.791) |
| **KPNC temporally distinct** | ≥ Moderate AS | 0.983<br>(0.980 – 0.985) | 0.823<br>(0.803 – 0.842) | 0.979<br>(0.977 – 0.981) | 0.762<br>(0.741 – 0.783) | 0.985<br>(0.984 – 0.987) | 0.791<br>(0.775 – 0.807) |
|  | Severe AS | 0.985<br>(0.981 – 0.988) | 0.731<br>(0.691 – 0.771) | 0.989<br>(0.988 – 0.991) | 0.636<br>(0.596 – 0.676) | 0.993<br>(0.992 – 0.994) | 0.680<br>(0.647 – 0.712) |
| **SHC** | ≥ Moderate AS | 0.989<br>(0.984 – 0.993) | 0.793<br>(0.740 – 0.843) | 0.991<br>(0.987 – 0.995) | 0.911<br>(0.872 – 0.948) | 0.977<br>(0.970 – 0.983) | 0.848<br>(0.811 – 0.882) |
|  | Severe AS | 0.985<br>(0.975 – 0.992) | 0.809<br>(0.723 – 0.885) | 0.985<br>(0.980 – 0.990) | 0.691<br>(0.600 – 0.777) | 0.992<br>(0.988 – 0.996) | 0.745<br>(0.673 – 0.809) |
| **CSMC** | ≥ Moderate AS | 0.978<br>(0.974 – 0.982) | 0.742<br>(0.702 – 0.781) | 0.980<br>(0.977 – 0.983) | 0.665<br>(0.624 – 0.706) | 0.986<br>(0.983 – 0.988) | 0.701<br>(0.667 – 0.733) |
|  | Severe AS | 0.989<br>(0.986 – 0.992) | 0.848<br>(0.797 – 0.895) | 0.988<br>(0.986 – 0.990) | 0.622<br>(0.565 – 0.678) | 0.996<br>(0.995 – 0.998) | 0.718<br>(0.670 – 0.763) |

AS, aortic stenosis; AUC, area under the receiver operating characteristic curve; PPV, positive predictive value; NPV, negative predictive value; KPNC, Kaiser Permanente Northern California; SHC, Stanford Healthcare; CSMC, Cedars-Sinai Medical Center; ≥ Moderate AS, moderate or severe AS

### Subgroup analysis

Analyzing patient subgroups further supported the generalizability of our framework. In the KPNC temporally distinct cohort, older patients (moderate or severe AS/severe AS: 0.983 [95% CI, 0.980–0.985] / 0.985 [0.981–0.988]), males (0.992 [0.984–0.998] / 0.998 [0.995–0.999]), females (0.980 [0.977–0.983] / 0.984 [0.978–0.988]), patients with obesity (BMI > 35: 0.985 [0.983–0.988] / 0.987 [0.983–0.990]), and those with reduced LVEF ≤ 50% (0.984 [0.982–0.987] / 0.986 [0.981–0.989]) all demonstrated high performance patients (0.881 [0.781–0.956] / 0.838 [0.622–0.995]). Performance was also robust in those with concomitant valvular disease such as AR, MR, TR, and MS. Slightly lower performance was observed in the small bicuspid aortic valve subgroup. Similar results were observed in the CSMC cohort: older patients (0.972 [0.966–0.976] / 0.986 [0.982–0.990]), males (0.979 [0.974–0.983] / 0.988 [0.983–0.992]), females (0.978 [0.971–0.983] / 0.991 [0.987–0.994]), obese patients (0.986 [0.978–0.993] / 0.994 [0.987–0.998]), and those with LVEF ≤ 50% (0.963 [0.952–0.973] / 0.977 [0.966–0.987]) consistently achieved high AUCs. Across both cohorts, severe AS detection generally outperformed moderate or severe AS detection, suggesting greater discriminative ability for severe cases (**Table 4**).

**Table 4.**
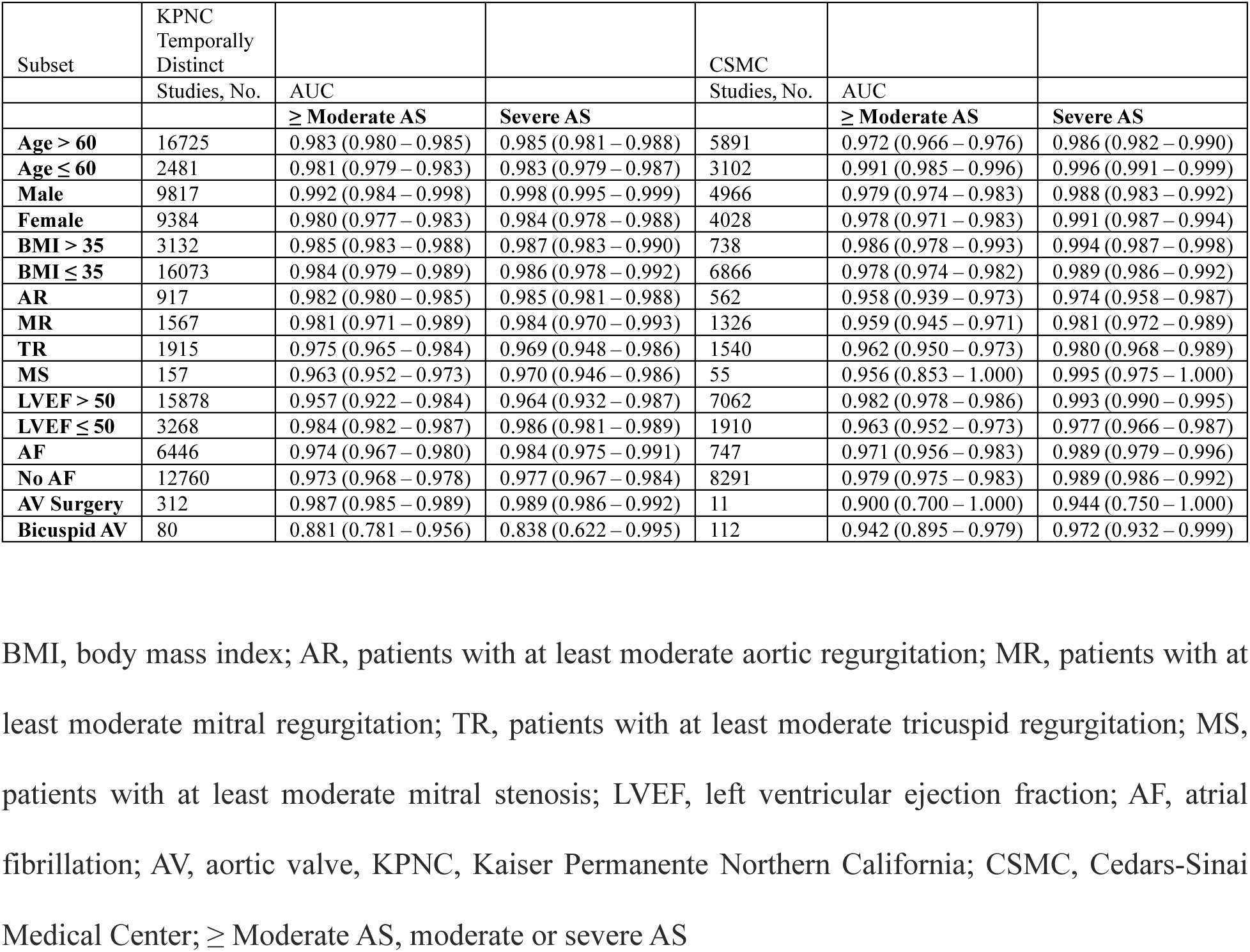
EchoNet-AS subset analysis.

| Subset | KPNC<br>Temporally<br>Distinct |  |  | CSMC |  |  |
| --- | --- | --- | --- | --- | --- | --- |
|  | Studies, No. | AUC |  | Studies, No. | AUC |  |
|  |  | **≥ Moderate AS** | **Severe AS** |  | **≥ Moderate AS** | **Severe AS** |
| **Age > 60** | 16725 | 0.983 (0.980 – 0.985) | 0.985 (0.981 – 0.988) | 5891 | 0.972 (0.966 – 0.976) | 0.986 (0.982 – 0.990) |
| **Age ≤ 60** | 2481 | 0.981 (0.979 – 0.983) | 0.983 (0.979 – 0.987) | 3102 | 0.991 (0.985 – 0.996) | 0.996 (0.991 – 0.999) |
| **Male** | 9817 | 0.992 (0.984 – 0.998) | 0.998 (0.995 – 0.999) | 4966 | 0.979 (0.974 – 0.983) | 0.988 (0.983 – 0.992) |
| **Female** | 9384 | 0.980 (0.977 – 0.983) | 0.984 (0.978 – 0.988) | 4028 | 0.978 (0.971 – 0.983) | 0.991 (0.987 – 0.994) |
| **BMI > 35** | 3132 | 0.985 (0.983 – 0.988) | 0.987 (0.983 – 0.990) | 738 | 0.986 (0.978 – 0.993) | 0.994 (0.987 – 0.998) |
| **BMI ≤ 35** | 16073 | 0.984 (0.979 – 0.989) | 0.986 (0.978 – 0.992) | 6866 | 0.978 (0.974 – 0.982) | 0.989 (0.986 – 0.992) |
| **AR** | 917 | 0.982 (0.980 – 0.985) | 0.985 (0.981 – 0.988) | 562 | 0.958 (0.939 – 0.973) | 0.974 (0.958 – 0.987) |
| **MR** | 1567 | 0.981 (0.971 – 0.989) | 0.984 (0.970 – 0.993) | 1326 | 0.959 (0.945 – 0.971) | 0.981 (0.972 – 0.989) |
| **TR** | 1915 | 0.975 (0.965 – 0.984) | 0.969 (0.948 – 0.986) | 1540 | 0.962 (0.950 – 0.973) | 0.980 (0.968 – 0.989) |
| **MS** | 157 | 0.963 (0.952 – 0.973) | 0.970 (0.946 – 0.986) | 55 | 0.956 (0.853 – 1.000) | 0.995 (0.975 – 1.000) |
| **LVEF > 50** | 15878 | 0.957 (0.922 – 0.984) | 0.964 (0.932 – 0.987) | 7062 | 0.982 (0.978 – 0.986) | 0.993 (0.990 – 0.995) |
| **LVEF ≤ 50** | 3268 | 0.984 (0.982 – 0.987) | 0.986 (0.981 – 0.989) | 1910 | 0.963 (0.952 – 0.973) | 0.977 (0.966 – 0.987) |
| **AF** | 6446 | 0.974 (0.967 – 0.980) | 0.984 (0.975 – 0.991) | 747 | 0.971 (0.956 – 0.983) | 0.989 (0.979 – 0.996) |
| **No AF** | 12760 | 0.973 (0.968 – 0.978) | 0.977 (0.967 – 0.984) | 8291 | 0.979 (0.975 – 0.983) | 0.989 (0.986 – 0.992) |
| **AV Surgery** | 312 | 0.987 (0.985 – 0.989) | 0.989 (0.986 – 0.992) | 11 | 0.900 (0.700 – 1.000) | 0.944 (0.750 – 1.000) |
| **Bicuspid AV** | 80 | 0.881 (0.781 – 0.956) | 0.838 (0.622 – 0.995) | 112 | 0.942 (0.895 – 0.979) | 0.972 (0.932 – 0.999) |

BMI, body mass index; AR, patients with at least moderate aortic regurgitation; MR, patients with at
least moderate mitral regurgitation; TR, patients with at least moderate tricuspid regurgitation; MS,
patients with at least moderate mitral stenosis; LVEF, left ventricular ejection fraction; AF, atrial
fibrillation; AV, aortic valve, KPNC, Kaiser Permanente Northern California; CSMC, Cedars-Sinai
Medical Center; ≥ Moderate AS, moderate or severe AS

### Explainability analysis

Saliency map analysis revealed that the view-specific models predominantly highlighted the aortic valve and surrounding regions on two-dimensional images, as well as the corresponding color Doppler signals around the aortic valve (**Figure 4A**). In the ensemble model without peak velocity, feature importance analysis demonstrated that features derived from the PLAX view contributed most strongly to the final prediction, followed by the apical and PSAX views. When peak aortic velocity was incorporated as an additional input, it became the single most influential feature, surpassing all view-specific model outputs, which was consistent with our expectations given its established role in assessing AS severity (**Figure 4B, C**).

**Figure 4.**
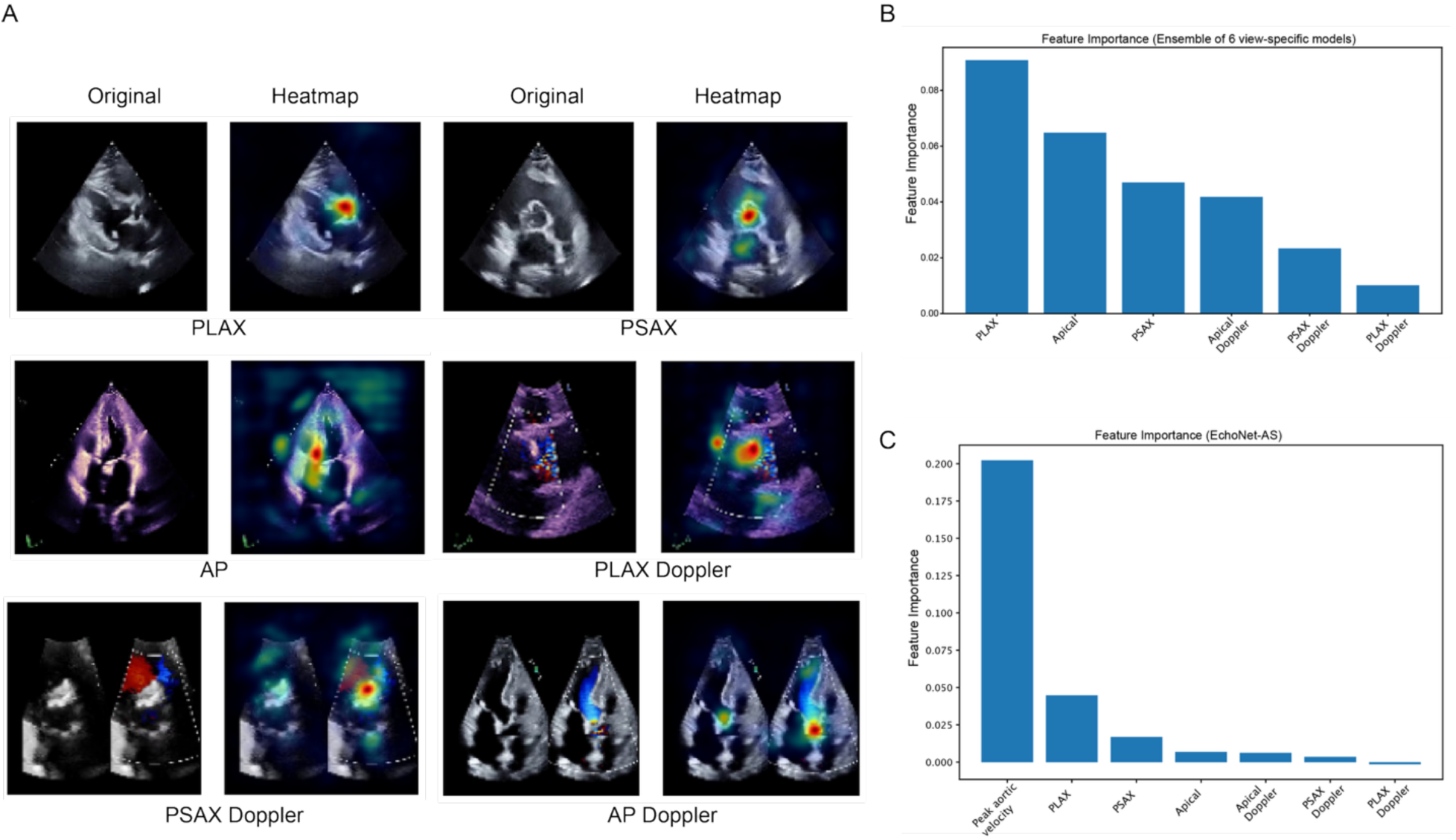
Model interpretability and feature importance analysis. A. Representative Smooth Grad visualizations for each of the six echocardiographic views (parasternal long axis [PLAX], parasternal short axis [PSAX], apical [AP], and their corresponding Doppler views) illustrating regions contributing most to model predictions of aortic stenosis (AS) severity. B, C. Model-level feature importance from the ensemble classifier. B shows feature importance for the ensemble of view-specific models. C shows feature importance when peak aortic velocity is included as an additional input feature.

## Discussion

In this study, we developed and validated EchoNet-AS, a novel automated deep learning– based framework that estimates AS severity by incorporating video-based information from multiple echocardiographic views along with automated Doppler measurements of peak aortic jet velocity. Our multi-view approach demonstrated excellent performance (AUC ranging from 0.964 to 0.989 for discriminating severe AS, and 0.955 – 0.978 for discriminating at least moderate AS), while maintaining favorable PPV/NPV, recall, specificity, and F1-scores across 4 independent test cohorts. This model consistently outperformed single-view models, underscoring the complementary value of multi-view assessment and quantitative hemodynamics. Although the absolute AUC gains compared to single-view models may appear modest, even small improvements in discrimination can translate into fewer misclassified patients in practice, potentially reducing unnecessary repeat echocardiograms, shortening delays in referral for valve intervention, and improving consistency across sites.

In recent years, AI have been applied to echocardiography across a range of tasks, including assessment of left ventricular systolic dysfunction^25,26^, detection of left ventricular hypertrophy and its various subtypes^27^, detection of valvular regurgitation^4–6,9^. For AS specifically, previous work has demonstrated the feasibility of predicting severe AS from a single PLAX view; for example, Dai et al. developed deep learning models to discriminate whether the mean transvalvular pressure is greater than 40 mmHg and AVA is smaller than 1 cm^2^ from PLAX videos, achieving an AUC of 0.79 in the held-out test dataset^10^. Holste et al. developed a deep learning model to predict severe AS from PLAX images using 17570 videos and demonstrated an AUC of 0.952 [0.941 – 0.963] on external validation datasets. Our PLAX view model showed similar or greater performance (AUC 0.919 – 0.972), and our work builds upon these studies by expanding the size of training dataset (210,193 videos from 16,076 studies), and systematically comparing single-view, multi-view, and view-plus-measurement strategies. EchoNet-AS generalized across an KPNC held-out cohort, a temporally distinct cohort from a later year, and two external cohort (SHC and CSMC) with differing patient and echocardiogram acquisition characteristics. We also observed some variability across sites and between tasks (severe AS vs moderate or severe AS). This may reflect differences in case mix and documentation practices, and the fact that the moderate–severe boundary is inherently more challenging and noisier, whereas clearly severe cases tend to be more distinct. This breadth of validation reduces the risk that performance reflects idiosyncrasies of a single site or period. Moreover, performance remained robust across subgroups defined by age, sex, and comorbidities, suggesting resilience to common clinical heterogeneity. Performance was somewhat lower in patients with bicuspid valves, likely reflecting different leaflet morphology and eccentric jets, but was similar across patients with preserved versus reduced LVEF and in those with concomitant valve disease. These findings suggest the framework is broadly applicable across typical clinical scenarios.

Consistent with current clinical practice, incorporating multiple views and Doppler information are necessary to assess AS severity. Multiple views capture complementary information about aortic valve morphology, jet orientation, and surrounding structures. Ensembling across views mitigates view-specific limitations (e.g., suboptimal probe angle, foreshortening, or shadowing) and stabilizes predictions. The importance of multi-view assessment is emphasized in the literature^14,28^. Beyond images, peak aortic jet velocity is a key quantitative marker of AS severity; incorporating an automated estimate anchors the model in physiological signal. The resulting framework therefore fuses structural (B-mode), flow (color Doppler), and quantitative (peak velocity) evidence—mirroring expert practice—and delivers consistent gains over image-only ensembles. By ensembling predictions from all six view-specific models, we achieved further improvements in classification performance, underscoring the complementary value of multi-view integration. The inclusion of quantitative measurement— specifically, peak aortic velocity—further enhanced predictive performance, which is consistent with its established clinical importance in AS grading. By flagging studies with severe or moderate AS, the system could help reduce the risk of missed severe AS on echocardiography and facilitate more timely intervention. In instances where the model’s prediction disagrees with the physician’s initial interpretation, a secondary review of the echocardiographic videos and reassessment of severity grading could be undertaken, which may further enhance diagnostic precision.

There are several limitations of our study. First, the ground truth for AS severity was based on clinical reports rather than a dedicated core lab analysis, which could introduce some variability in the reference standard. However, this approach reflects real-world clinical practice, and all studies were interpreted by board-certified cardiologists following standardized guidelines. Second, while peak velocity is a major AS severity anchor, future work could incorporate additional quantitative measures (e.g., mean gradient or aortic valve area^14^) and other clinical parameters. Finally, this study lacks the prospective validation; future work through prospective randomized and integrated into routine workflow^26,29,30^, is required to establish effectiveness and the impact on patient care.

## Conclusions

In summary, we propose EchoNet-AS, an open source, video-based framework that integrates multi-view B-modes, Doppler color flow and an automated Doppler peak-velocity measurement to estimate AS severity. Validated across four independent cohorts, EchoNet-AS demonstrated robust performance, particularly for detecting severe AS, and could serve as a valuable clinical decision support tool for AS assessment in routine clinical practice.

## Declarations

### Data availability

The dataset of echocardiography videos and reports used to train EchoNet-AS is not available for public sharing, given the restrictions in our institutional review board approval.

### Code availability

Our code and model weights are available at https://github.com/echonet/AS.

### Ethical approval

This study received approval from the Institutional Review Boards of KPNC, SHC and CSMC. Informed consent was waived due to the study’s use of secondary analysis of existing data.

---

**sTable 1.** Model performance to detect severe or at least moderate aortic stenosis.

| Cohort or class | AUC | PPV | NPV | Recall | Specificity | F1 |
| --- | --- | --- | --- | --- | --- | --- |
| **Internal Test** |  |  |  |  |  |  |
| ≥ Moderate AS | 0.955 (0.946 – 0.964) | 0.842 (0.811 – 0.872) | 0.909 (0.892 – 0.926) | 0.828 (0.796 – 0.859) | 0.917 (0.900 – 0.933) | 0.835 (0.811 – 0.858) |
| Severe AS | 0.964 (0.952 – 0.973) | 0.763 (0.701 – 0.821) | 0.960 (0.949 – 0.970) | 0.729 (0.668 – 0.789) | 0.966 (0.956 – 0.975) | 0.746 (0.696 – 0.791) |
| **Temporal Test** |  |  |  |  |  |  |
| ≥ Moderate AS | 0.983 (0.980 – 0.985) | 0.823 (0.803 – 0.842) | 0.979 (0.977 – 0.981) | 0.762 (0.741 – 0.783) | 0.985 (0.984 – 0.987) | 0.791 (0.775 – 0.807) |
| Severe AS | 0.985 (0.981 – 0.988) | 0.731 (0.691 – 0.771) | 0.989 (0.988 – 0.991) | 0.636 (0.596 – 0.676) | 0.993 (0.992 – 0.994) | 0.680 (0.647 – 0.712) |
| **SHC** |  |  |  |  |  |  |
| ≥ Moderate AS | 0.989 (0.984 – 0.993) | 0.793 (0.740 – 0.843) | 0.991 (0.987 – 0.995) | 0.911 (0.872 – 0.948) | 0.977 (0.970 – 0.983) | 0.848 (0.811 – 0.882) |
| Severe AS | 0.985 (0.975 – 0.992) | 0.809 (0.723 – 0.885) | 0.985 (0.980 – 0.990) | 0.691 (0.600 – 0.777) | 0.992 (0.988 – 0.996) | 0.745 (0.673 – 0.809) |
| **CSMC** |  |  |  |  |  |  |
| ≥ Moderate AS | 0.978 (0.974 – 0.982) | 0.742 (0.702 – 0.781) | 0.980 (0.977 – 0.983) | 0.665 (0.624 – 0.706) | 0.986 (0.983 – 0.988) | 0.701 (0.667 – 0.733) |
| Severe AS | 0.989 (0.986 – 0.992) | 0.848 (0.797 – 0.895) | 0.988 (0.986 – 0.990) | 0.622 (0.565 – 0.678) | 0.996 (0.995 – 0.998) | 0.718 (0.670 – 0.763) |

Performance metrics with 95% confidence intervals for the prediction of “severe AS” and “moderate or severe AS” across four cohorts. AUC, area under the receiver operating characteristic curve; PPV, positive predictive value, NPV, negative predictive value; ≥ Moderate AS, moderate or severe AS

**sTable 2.** Severe AS detection performance in CSMC replicated cohort.

|  |  |
| --- | --- |
| Models |  |
|  | CSMC replicated cohort<br>(N=4226) |
| **EchoNet-AS** | 0.972<br>(0.963 – 0.979) |
| Ensemble of B Modes + Color Dopplers | 0.971<br>(0.961 – 0.978) |
| Ensemble of B Modes | 0.968<br>(0.958 – 0.976) |
| PLAX | 0.956<br>(0.940 – 0.969) |
| PLAX (Yale model, value from published<br>paper<sup>11</sup>) | 0.952<br>(0.941 – 0.963) |

## References

1. Okumus, N., Abraham, S., Puri, R. & Tang, W. H. W. Aortic valve disease, transcatheter aortic valve replacement, and the heart failure patient: A state-of-the-art review. JACC Heart Fail. 11, 1070–1083 (2023).

2. Eugène, M. et al. Contemporary management of severe symptomatic aortic stenosis. J. Am. Coll. Cardiol. 78, 2131–2143 (2021).

3. Tan, M. C. et al. Trends and disparities in valvular heart disease mortality in the United States from 1999 to 2020. J. Am. Heart Assoc. 13, e030895 (2024).

4. Vrudhula, A. et al. High-throughput deep learning detection of mitral regurgitation. Circulation 150, 923–933 (2024).

5. Binder, C. et al. Automated aortic regurgitation detection and quantification: A deep learning approach using multi-view echocardiography. medRxiv (2025) doi:10.1101/2025.03.18.25323918.

6. Vrudhula, A. et al. Automated deep learning phenotyping of tricuspid regurgitation in echocardiography. JAMA Cardiol. 10, 595–602 (2025).

7. Christensen, M., Vukadinovic, M., Yuan, N. & Ouyang, D. Vision-language foundation model for echocardiogram interpretation. Nat. Med. 30, 1481–1488 (2024).

8. Vukadinovic, M. et al. EchoPrime: A multi-video view-informed vision-language model for comprehensive echocardiography interpretation. arXiv [cs.CV] (2024).

9. Long, A. et al. Deep learning for echocardiographic assessment and risk stratification of aortic, mitral, and tricuspid regurgitation: the DELINEATE-regurgitation study. Eur. Heart J. 46, 2780–2791 (2025).

10. Dai, W., Nazzari, H., Namasivayam, M., Hung, J. & Stultz, C. M. Identifying aortic stenosis with a single parasternal long-axis video using deep learning. J. Am. Soc. Echocardiogr. 36, 116–118 (2023).

11. Holste, G. et al. Severe aortic stenosis detection by deep learning applied to echocardiography. Eur. Heart J. 44, 4592–4604 (2023).

12. Krishna, H. et al. Fully automated artificial intelligence assessment of aortic stenosis by echocardiography. J. Am. Soc. Echocardiogr. 36, 769–777 (2023).

13. Park, J., Kim, J., Jeon, J. & Yoon, Y. E. Utilizing deep learning for accurate assessment of aortic valve stenosis: case series for clinical applications. J. Cardiovasc. Imaging 33, 3 (2025).

14. Baumgartner, H. et al. Recommendations on the echocardiographic assessment of aortic valve stenosis: A focused update from the European Association of cardiovascular imaging and the American Society of Echocardiography. J. Am. Soc. Echocardiogr. 30, 372–392 (2017).

15. Vahanian, A. et al. 2021 ESC/EACTS Guidelines for the management of valvular heart disease. Eur. Heart J. 43, 561–632 (2022).

16. Baumgartner, H. et al. Echocardiographic assessment of valve stenosis: EAE/ASE recommendations for clinical practice. J. Am. Soc. Echocardiogr. 22, 1–23; quiz 101–2 (2009).

17. Mitchell, C. et al. Guidelines for performing a comprehensive transthoracic echocardiographic examination in adults: Recommendations from the American society of echocardiography. J. Am. Soc. Echocardiogr. 32, 1–64 (2019).

18. Tran, D. et al. A Closer Look at Spatiotemporal Convolutions for Action Recognition. in 2018 IEEE/CVF Conference on Computer Vision and Pattern Recognition 6450–6459 (IEEE, 2018).

19. Pytorch-Lightning: Pretrain, Finetune ANY AI Model of ANY Size on Multiple GPUs, TPUs with Zero Code Changes. (Github).

20. Defazio, A. et al. The Road Less Scheduled. in Advances in Neural Information Processing Systems (eds. Globerson, A. et al.) vol. 37 9974–10007 (Curran Associates, Inc., 2024).

21. Micikevicius, P., et al. Mixed Precision Training. arXiv [cs.AI] (2017).

22. Sahashi, Y. et al. Artificial intelligence automation of echocardiographic measurements. medRxiv 2025.03.18.25324215 (2025) doi:10.1101/2025.03.18.25324215.

23. Pedregosa, F. et al. Scikit-learn: Machine Learning in Python. J. Mach. Learn. Res. 12, 2825–2830 (2011).

24. Smilkov, D., Thorat, N., Kim, B., Viégas, F. & Wattenberg, M. SmoothGrad: removing noise by adding noise. arXiv [cs.LG] (2017).

25. Ouyang, D. et al. Video-based AI for beat-to-beat assessment of cardiac function. Nature 580, 252–256 (2020).

26. He, B. et al. Blinded, randomized trial of sonographer versus AI cardiac function assessment. Nature 616, 520–524 (2023).

27. Duffy, G. et al. High-throughput precision phenotyping of left ventricular hypertrophy with cardiovascular deep learning. JAMA Cardiol. 7, 386–395 (2022).

28. Thaden, J. J., Nkomo, V. T., Lee, K. J. & Oh, J. K. Doppler imaging in aortic stenosis: The importance of the nonapical imaging windows to determine severity in a contemporary cohort. J. Am. Soc. Echocardiogr. 28, 780–785 (2015).

29. Yao, X. et al. Artificial intelligence-enabled electrocardiograms for identification of patients with low ejection fraction: a pragmatic, randomized clinical trial. Nat. Med. 27, 815–819 (2021).

30. Sakamoto, A. et al. Artificial Intelligence-based Automated Echocardiographic Analysis and the Workflow of Sonographers: A randomized crossover trial. medRxiv 2025.08.20.25334115 (2025) doi:10.1101/2025.08.20.25334115.

